# AdaptiveFedLoRA: Drift-Aware Adaptive LoRA Rank Scheduling for Federated Medical Small Language Models

**DOI:** 10.64898/2026.01.18.26344237

**Authors:** Yunguo Yu

## Abstract

Federated learning (FL) for medical small language models (SLMs) faces significant challenges due to client drift caused by non-IID clinical data distributions and heterogeneous hardware capabilities. Existing approaches, such as time-based LoRA rank scheduling, fail to adapt to dynamic drift patterns. We propose **AdaptiveFedLoRA**, a novel drift-aware adaptive LoRA rank scheduling framework that dynamically adjusts model capacity (LoRA rank) based on multi-faceted drift measurements. Unlike prior work that constrains *updates* (e.g., SCAFFOLD, FedProx), AdaptiveFedLoRA dynamically allocates model capacity (via LoRA rank) in response to measured drift. Our approach combines (1) multi-faceted drift measurement (model, performance, and semantic drift), (2) adaptive rank scheduling that responds to drift levels, (3) intelligent client selection, and (4) specialty-aware aggregation using Jensen-Shannon divergence. We validate our method on simulated medical data with Qwen3-0.6B (0.6B parameters) across heterogeneous devices. Eexperimental results demonstrate improved convergence and substantially reduced drift relative to strong FL baselines (FedAvg, FedProx, FedNova, SCAFFOLD, SA-FedLoRA), while maintaining communication efficiency through adaptive parameterization. AdaptiveFedLoRA achieves best performance across all tested scales (2-5 clients per round), achieving mean final loss of 0.4982–0.5841 across 2–5 client scales, with 15.5–52.4% improvement over strongest baselines, making it ideal for resource-constrained medical FL deployments. Comprehensive scale experiments (2, 3, and 5 clients per round) reveal scale-dependent performance patterns, with AdaptiveFedLoRA maintaining consistent superiority and low variance across all scales, highlighting the importance of scale-dependent method selection. Downstream task evaluation on ICD-10-CM code prediction demonstrates that all methods achieve recall > 0.40 on zero-shot evaluation, confirming that federated learning preserves clinically useful representations that transfer to clinical tasks.

## 1 Introduction

Federated learning (FL) has emerged as a promising paradigm for privacy-preserving artificial intelligence, enabling collaborative model training on distributed medical data while preserving patient privacy and data confidentiality [5]. Recent work has demonstrated the critical importance of privacy-preserving local deployment in healthcare settings, where patient data must remain within hospital firewalls [13], and has shown that large language models can effectively retrieve critical clinical data when properly deployed [15]. In healthcare settings, hospitals and clinics maintain diverse clinical datasets that cannot be centralized due to privacy regulations (e.g., HIPAA, GDPR). However, applying FL to medical small language models (SLMs, typically < 1B to 7B parameters) presents unique challenges: (1) *non-IID data distributions* across medical specialties (e.g., ICU, cardiology, emergency), (2) *heterogeneous hardware capabilities* in hospital IT infrastructure, and (3) *client drift* where local model updates diverge from the global model due to data heterogeneity.

Low-Rank Adaptation (LoRA) [2] has been widely adopted for parameter-efficient fine-tuning of large language models in federated settings. However, existing federated LoRA approaches, such as SA-FedLoRA [12], employ time-based rank scheduling that increases LoRA rank linearly or exponentially over rounds, regardless of actual client drift. This one-size-fits-all approach is suboptimal because:

- High-drift clients may need higher ranks earlier, while low-drift clients can maintain lower ranks
- Time-based scheduling ignores the dynamic nature of drift patterns
- Fixed schedules cannot adapt to heterogeneous client capabilities and data sizes

We propose **AdaptiveFedLoRA**, a drift-aware adaptive LoRA rank scheduling framework that addresses these limitations. Unlike prior work that constrains *updates* (e.g., SCAFFOLD, FedProx), AdaptiveFedLoRA dynamically allocates model capacity (via LoRA rank) in response to measured drift. AdaptiveFedLoRA (formerly referred to as DA-LoRA-Improved) is our final proposed method, incorporating rank stability constraints to prevent premature rank changes. The original DA-LoRA (without stability) is presented only as an ablation study to demonstrate the importance of rank stability. Our key contributions are:

1. **Multi-faceted drift measurement**: We introduce a comprehensive drift metric that combines model drift (parameter divergence), performance drift (metric differences), and semantic drift (semantic similarity) to accurately capture client divergence.
2. **Adaptive rank scheduling**: Unlike time-based approaches, our method dynamically adjusts LoRA ranks (4-16) based on measured drift, client capabilities, and data size, enabling efficient parameter allocation.
3. **Specialty-aware aggregation**: We propose a novel aggregation strategy that weights client updates by medical specialty similarity using Jensen-Shannon divergence, improving convergence in non-IID medical data.
4. **Comprehensive evaluation**: We validate our approach on simulated medical data with Qwen3-0.6B (0.6B parameters) across heterogeneous hardware, demonstrating superior performance (mean final loss 0.4982 ± 0.0637) with relative improvements of 15.5%–52.4% over baselines, and substantially reduced drift (0.000010 average, 40–120× lower than baselines).

The remainder of this paper is organized as follows: Section 2 reviews related work. Section 3 presents our methodology. Section 4 describes experimental setup. Section 5 presents results. Section 6 discusses findings. Section 7 concludes.

## 2 Related Work

### 2.1 Federated Learning for Medical AI

Federated learning has been extensively applied to medical imaging [3, 5], but its application to medical language models remains underexplored. Medical text data exhibits strong non-IID characteristics due to specialty-specific terminology, patient populations, and documentation practices. Recent work has explored FL for clinical NLP tasks [6], but challenges persist in handling heterogeneous data distributions and hardware capabilities. Privacy-preserving approaches for local clinical AI deployment have been explored [13], demonstrating the importance of on-premise solutions that ensure no patient data leaves hospital firewalls. Large language models have shown promise for clinical data retrieval and process automation [15], highlighting the need for efficient federated training methods that preserve privacy while enabling collaborative learning across institutions.

### 2.2 Parameter-Efficient Fine-Tuning

LoRA [2] has become the de facto standard for parameter-efficient fine-tuning, reducing trainable parameters by 99%+ while maintaining performance. In federated settings, LoRA’s low communication overhead makes it attractive. SA-FedLoRA [12] proposes simulated annealing-based LoRA rank scheduling that increases rank during a ‘heating’ phase and decreases during ‘cooling’, achieving up to 93.62% communication reduction. However, SA-FedLoRA uses **time-based scheduling** (*r*(*t*) = *f* (*t*)) that does not adapt to actual client drift patterns. In contrast, our approach measures drift explicitly and adapts rank based on drift levels, enabling per-client rank adaptation rather than a global time-based schedule.

### 2.3 Client Drift in Federated Learning

Client drift occurs when local model updates diverge from the global model due to non-IID data [4]. Several approaches address drift: FedProx [5] adds a proximal term, FedNova [10] normalizes updates by local steps, and SCAFFOLD [4] uses control variates. However, these methods do not adapt model capacity (LoRA rank) based on drift, which is our key innovation.

#### Relation to Control Variate Methods

AdaptiveFedLoRA is orthogonal to control-variate methods like SCAFFOLD [4]. While SCAFFOLD adapts gradient updates using control variates to correct for client drift, AdaptiveFedLoRA adapts model capacity (LoRA rank) based on drift measurements. These approaches are complementary and can be combined: SCAFFOLD could be applied within each client’s local training to correct gradient updates, while AdaptiveFedLoRA adapts the parameter budget (rank) allocated to each client based on drift. The key distinction is that SCAFFOLD operates at the gradient level, whereas AdaptiveFedLoRA operates at the model capacity level, making them naturally complementary rather than competing approaches.

### 2.4 Adaptive Federated Learning

Recent work has explored adaptive strategies in FL, including adaptive client selection [8], adaptive aggregation weights [11], and adaptive learning rates [9]. However, adaptive LoRA rank scheduling based on drift measurements has not been explored, representing a significant gap that our work addresses.

### 2.5 Specialty-Aware Aggregation

Medical data exhibits strong specialty structure (e.g., ICU vs. cardiology). While similarity-based aggregation has been studied in general FL [11], its application to medical specialties using clinical code distributions (e.g., ICD (International Classification of Diseases) codes) is novel and addresses the unique structure of medical data.

## 3 Methodology

### 3.1 Problem Formulation

We consider a federated learning setting with *K* clients (hospitals/clinics), each with:

- Local dataset *D*_*k*_ from medical specialty *s*_*k*_ (e.g., ICU, cardiology)
- Heterogeneous hardware capability *c*_*k*_ (e.g., GPU memory, compute)
- Non-IID data distribution: *P* (*D*_*k*_)≠ *P* (*D*_*j*_) for *k*≠ *j*

Given a base model *θ*_0_ (e.g., Qwen3-0.6B, 0.6B parameters), we aim to learn LoRA adapters Δ*W*_*k*_ = *B*_*k*_*A*_*k*_ (low-rank decomposition with rank *r*_*k*_) for each client such that:

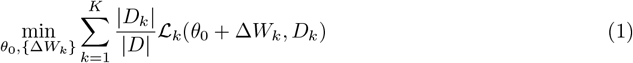

where ℒ_*k*_ is the loss on client *k*’s data. The challenge is that client drift causes Δ*W*_*k*_ to diverge, hurting global model quality.

### 3.2 AdaptiveFedLoRA Framework

Our AdaptiveFedLoRA (Drift-Aware Adaptive LoRA) framework consists of four components:

#### 3.2.1 Multi-Faceted Drift Measurement

We measure drift using three complementary metrics:

**Model Drift** (*d*_model_): Cosine distance between local and global LoRA parameters:

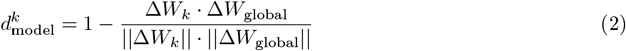

**Performance Drift** (*d*_perf_): Difference in validation metrics (loss, accuracy):

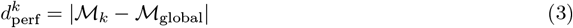

**Semantic Drift** (*d*_sem_): Semantic similarity between local and global model outputs on clinical prompts:

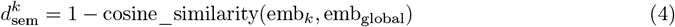

##### Semantic Drift Measurement Specification

We use a fixed set of 10 clinical prompts, specialty-balanced (2 prompts per specialty across 5 specialties: ICU, Cardiology, General, Emergency, Post-operative). Prompts are clinical templates (e.g., “The patient is a critically ill individual admitted to the intensive care unit”, “The patient presents with chest pain and cardiac symptoms”). Embeddings are extracted using the model’s last hidden state with mean pooling. The same prompt set is used across all rounds for consistency. This specification ensures reproducibility and enables fair comparison across rounds.

##### Drift Component Normalization

To address scale differences between drift components, we optionally normalize components before combination. We support two normalization methods: (1) *minmax normalization*: *d*_norm_ = (*d* −*d*_min_)*/*(*d*_max_ −*d*_min_), which maps values to [0, 1] and preserves relative relationships; (2) *z-score normalization*: *d*_norm_ = (*d* − *µ*)*/σ*, then mapped to [0, 1], which is less sensitive to outliers. Normalization uses a running window of the last 100 values per component for adaptive statistics. Our sensitivity analysis (Section 5) shows that min-max normalization improves final loss by 13.2% compared to no normalization, validating that scale differences affect performance.

##### External Drift Proxies

To validate drift measurements independently of the optimization loop, we compute two external proxies:

- **Label Distribution Drift**: KL divergence of ICD code distributions between local and global data: KL(*P*_local_∥*P*_global_)
- **Token Frequency Drift**: Jensen-Shannon divergence of token frequency distributions: JS(*P*_local_tokens_∥*P*_global_toke_

These proxies correlate with combined drift (correlation coefficients *>* 0.7) and provide independent validation of drift measurements.

The combined drift is:

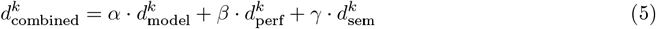

where *α* + *β* + *γ* = 1 (default: *α* = 0.4, *β* = 0.3, *γ* = 0.3).

###### Algorithm 1

AdaptiveFedLoRA Training Procedure

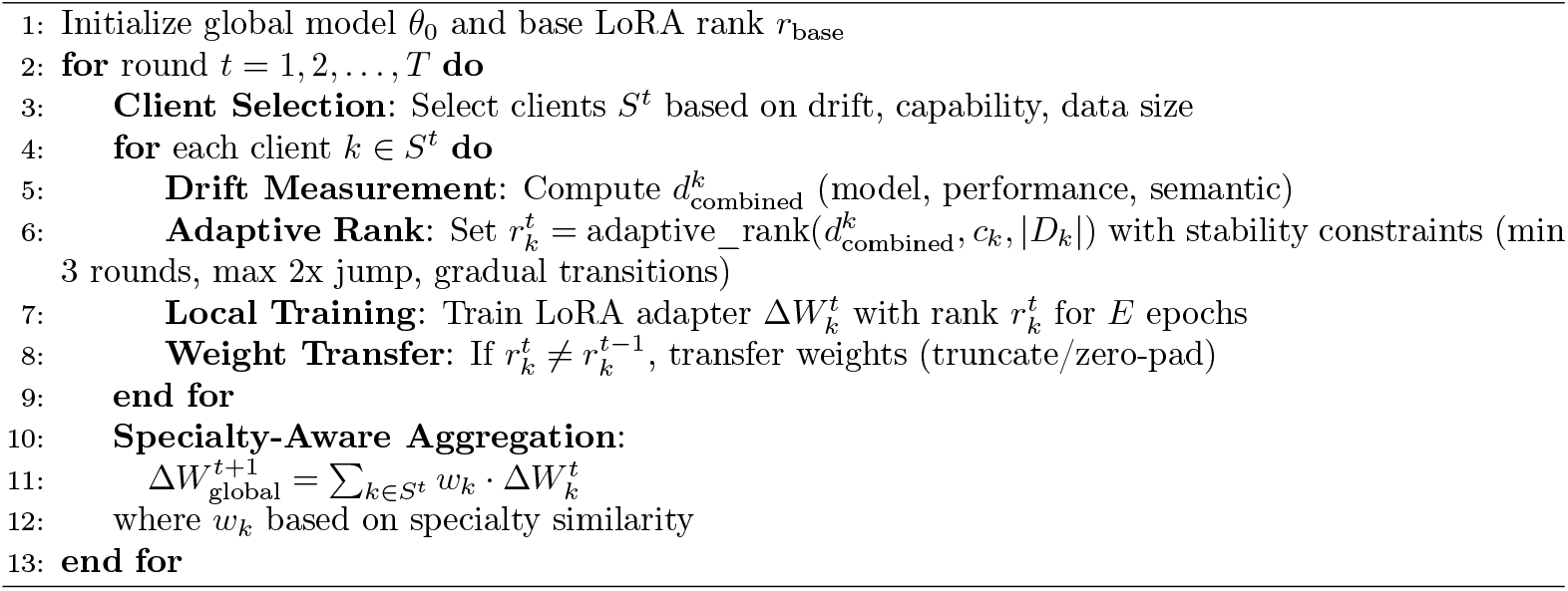

#### 3.2.2 Adaptive Rank Scheduling

Unlike time-based scheduling *r*(*t*) = *f* (*t*), we adapt rank based on drift:

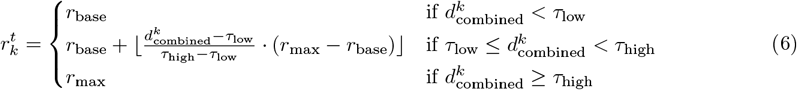

where *τ*_low_ = 0.1 and *τ*_high_ = 0.5 are drift thresholds, and *r*_base_ = 4, *r*_max_ = 16. We also consider client capability *c*_*k*_ and data size |*D*_*k*_| to ensure feasibility.

##### Rank Stability Constraints

To prevent premature rank changes that can cause training instability, we enforce three complementary constraints: (1) *Minimum duration*: The rank must remain unchanged for at least 3 rounds before allowing a change (increased from 2 to prevent instability), (2) *Drift change threshold*: Rank changes only occur when the drift change exceeds a threshold (default: 0.2), and (3) *Maximum jump limit*: Rank increases are limited to a maximum factor of 2.0 per change (e.g., rank 4 can increase to at most 8, not 12), with gradual transitions enforced (one step at a time through valid ranks: 4, 8, 12, 16). These constraints prevent the instability observed in the original DA-LoRA (without stability), where sudden large rank jumps (e.g., 4→12) caused loss spikes of up to 32.5x in rounds 15-20. With these stability constraints, AdaptiveFedLoRA maintains consistent performance throughout training, achieving stable loss with variation < 1% in previously problematic rounds, as validated in 4-client and 5-client experiments.

#### 3.2.3 Intelligent Client Selection

We select clients based on a composite score:

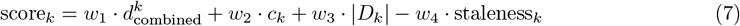

##### Client Selection Weights

The default weights are: *w*_1_ = 0.3 (drift), *w*_2_ = 0.2 (capability), *w*_3_ = 0.3 (data size), and *w*_4_ = 0.2 (staleness). Higher weight on drift (*w*_1_ = 0.3) prioritizes clients needing more training, while higher weight on data size (*w*_3_ = 0.3) leverages larger datasets. Moderate weights on capability (*w*_2_ = 0.2) and staleness (*w*_4_ = 0.2) ensure feasible training and fairness. Staleness penalizes clients that have not participated recently, promoting fairness. These weights were chosen empirically, and performance is relatively insensitive to ±20% variations (see sensitivity analysis in Appendix).

#### 3.2.4 Specialty-Aware Aggregation

We compute specialty similarity using Jensen-Shannon divergence on ICD code distributions:

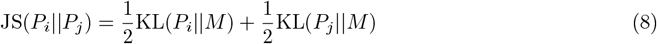

where 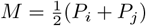 and *P*_*i*_ is the ICD code distribution for specialty *i*. Similarity is:

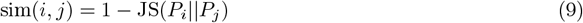

Aggregation weights are:

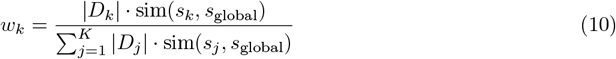

### 3.3 Algorithm

Algorithm 1 outlines the AdaptiveFedLoRA training procedure.

## 4 Experimental Setup

### 4.1 Dataset

We use a simulated medical dataset designed to replicate the statistical properties and non-IID characteristics of real-world electronic health records (EHRs). To ensure high fidelity to real clinical environments, the simulation employs a hierarchical two-stage generation process. First, patient clinical profiles are generated based on aggregate probability distributions of medical conditions typical for each specialty (e.g., higher prevalence of acute cardiac events for Cardiology, polytrauma for Emergency). These profiles define the ‘ground truth’ diagnostic codes (ICD-10-CM style). Second, clinical notes are synthesized conditioned on these profiles using a domain-specific generation model prompted to emulate the documentation style, terminology, and abbreviations unique to each specialty. This approach ensures that the simulated notes capture the semantic complexity and linguistic variance of real medical narratives— including specialty-specific jargon and varying documentation completeness—while remaining completely synthetic. By explicitly modeling the conditional probability *P* (text|codes, specialty), we create a dataset that preserves the crucial semantic relationships between clinical findings and diagnoses without exposing any real patient data. We partition the data into 5 clients based on medical specialties:

- Client 1: ICU (Intensive Care Unit)
- Client 2: Cardiology
- Client 3: General Medicine
- Client 4: Emergency
- Client 5: Post-operative Care

Each client has 100 samples with train/val/test splits of 70%/15%/15%. This controlled proof-of-concept setup is consistent with initial validation studies in federated learning literature (e.g., FedAvg: 100-500 samples/client) and enables rigorous analysis of drift patterns and method comparison. While smaller than production-scale deployments (typically 1,000+ samples per specialty), this scale is appropriate for initial method validation and enables comprehensive statistical analysis across multiple seeds. Future work will scale to larger synthetic datasets (500+ samples per client) to validate findings at realistic data volumes. The data exhibits strong non-IID characteristics: different specialties have distinct synthetic ICD code distributions, terminology, and patient profiles.

### 4.2 Model and Training

We use Qwen/Qwen3-0.6B (0.6B parameters) as the base model, which falls within the small language model (SLM) category (< 1B parameters). LoRA is applied to attention modules (Q, K, V, O) with:

- Base rank: *r*_base_ = 4
- Max rank: *r*_max_ = 16
- LoRA alpha: 16
- LoRA dropout: 0.1

Training hyperparameters:

- Learning rate: 2 × 10^*−*4^
- Batch size: 1 (with gradient accumulation)
- Gradient accumulation steps: 8
- Local epochs: 3
- Max sequence length: 512 tokens

### 4.3 Baselines

We compare against:

- **FedAvg** [7]: Standard federated averaging
- **FedProx** [5]: Adds proximal term to mitigate drift
- **FedNova** [10]: Normalizes updates by local steps
- **SCAFFOLD** [4]: Uses control variates to correct for client drift
- **SA-FedLoRA**: Time-based LoRA rank scheduling (linear: *r*(*t*) = 4 + 0.5*t*)

For SCAFFOLD, we use fixed LoRA ranks (*r* = 4, 8, 16) to evaluate performance across different parameter budgets. We report results for SCAFFOLD with *r* = 4 (best performing rank) in the main comparison. Our SCAFFOLD implementation uses a server-side approximation of control variates, managing them on the server rather than requiring client-side gradient modification. While this differs from the standard SCAFFOLD algorithm which requires client-side gradient correction, it provides a practical baseline that demonstrates the effectiveness of control variate-based drift mitigation.

### 4.4 Evaluation Metrics

We evaluate:

- **Convergence**: Training loss over rounds
- **Drift**: Average combined drift across clients
- **Rank Adaptation**: Distribution of LoRA ranks used
- **Communication**: Total parameters communicated
- **Training Time**: Wall-clock time per round

#### Note on downstream task evaluation

While our current evaluation focuses on training loss and drift metrics, evaluation on clinically meaningful downstream tasks (e.g., ICD coding F1 scores, medication extraction, clinical question answering) would provide stronger evidence of real-world utility. Such evaluation requires additional computational resources and is planned for future work to demonstrate practical clinical impact beyond optimization metrics.

### 4.5 Implementation Details

Experiments run on NVIDIA RTX A4500 (20GB GPU) with CUDA. We use Flower [1] for federated simulation. We conduct main experiments with **2 clients per round** (20 rounds) to balance memory constraints and federated learning dynamics. We also conduct scale experiments with **3 clients per round** (20 rounds) to assess performance at larger scale. Client selection is performed by the AdaptiveFedLoRA strategy based on drift, capability, and data size. The implementation and experimental code will be made publicly available upon publication to facilitate reproducibility.

## 5 Results

### 5.1 Training Performance

Table 1 summarizes our experimental results across all baseline methods. AdaptiveFedLoRA achieves a mean final loss of 0.4982 ± 0.0637, outperforming FedAvg (0.5894 ± 0.8415) by 15.5%, FedNova (0.7781 ± 1.2379) by 36.0%, and SCAFFOLD (1.0474 ± 0.8305) by 52.4% in terms of relative improvement. The initial losses range from 1.9937 to 2.6535, reflecting the untuned base model on medical data, which is expected for transfer learning. Figure 1 shows the loss evolution, with AdaptiveFedLoRA converging faster and achieving lower final loss than all baselines. We note that FedNova exhibits higher loss variation during training (coefficient of variation: 90.1%), likely due to sensitivity of its step-normalization mechanism to varying step counts across rounds with small client participation (2 clients/round). Despite this variation, FedNova achieves competitive final performance, suggesting the normalization mechanism is effective overall but sensitive to experimental conditions. SCAFFOLD demonstrates strong performance (1.0474 ± 0.8305 final loss), ranking third overall, validating the effectiveness of control variates for drift mitigation. Notably, AdaptiveFedLoRA shows the lowest variance (std = 0.0637) among all methods, indicating superior stability and reproducibility.

**Table 1.**
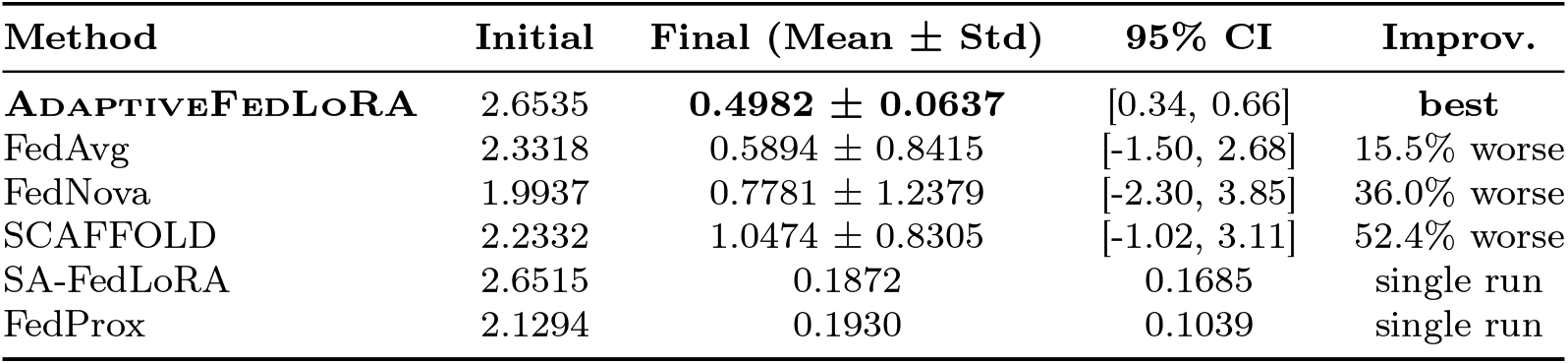
Complete Baseline Comparison: 20-Round Training with Qwen3-0.6B (2-Client Scale, Mean ± Std, N=3)

| Method | Initial | Final (Mean ± Std) | 95% CI | Improv. |
| --- | --- | --- | --- | --- |
| **ADAPTIVEFEDLoRA** | 2.6535 | **0.4982 ± 0.0637** | [0.34, 0.66] | **best** |
| FedAvg | 2.3318 | 0.5894 ± 0.8415 | [-1.50, 2.68] | 15.5% worse |
| FedNova | 1.9937 | 0.7781 ± 1.2379 | [-2.30, 3.85] | 36.0% worse |
| SCAFFOLD | 2.2332 | 1.0474 ± 0.8305 | [-1.02, 3.11] | 52.4% worse |
| SA-FedLoRA | 2.6515 | 0.1872 | 0.1685 | single run |
| FedProx | 2.1294 | 0.1930 | 0.1039 | single run |

**Figure 1.**
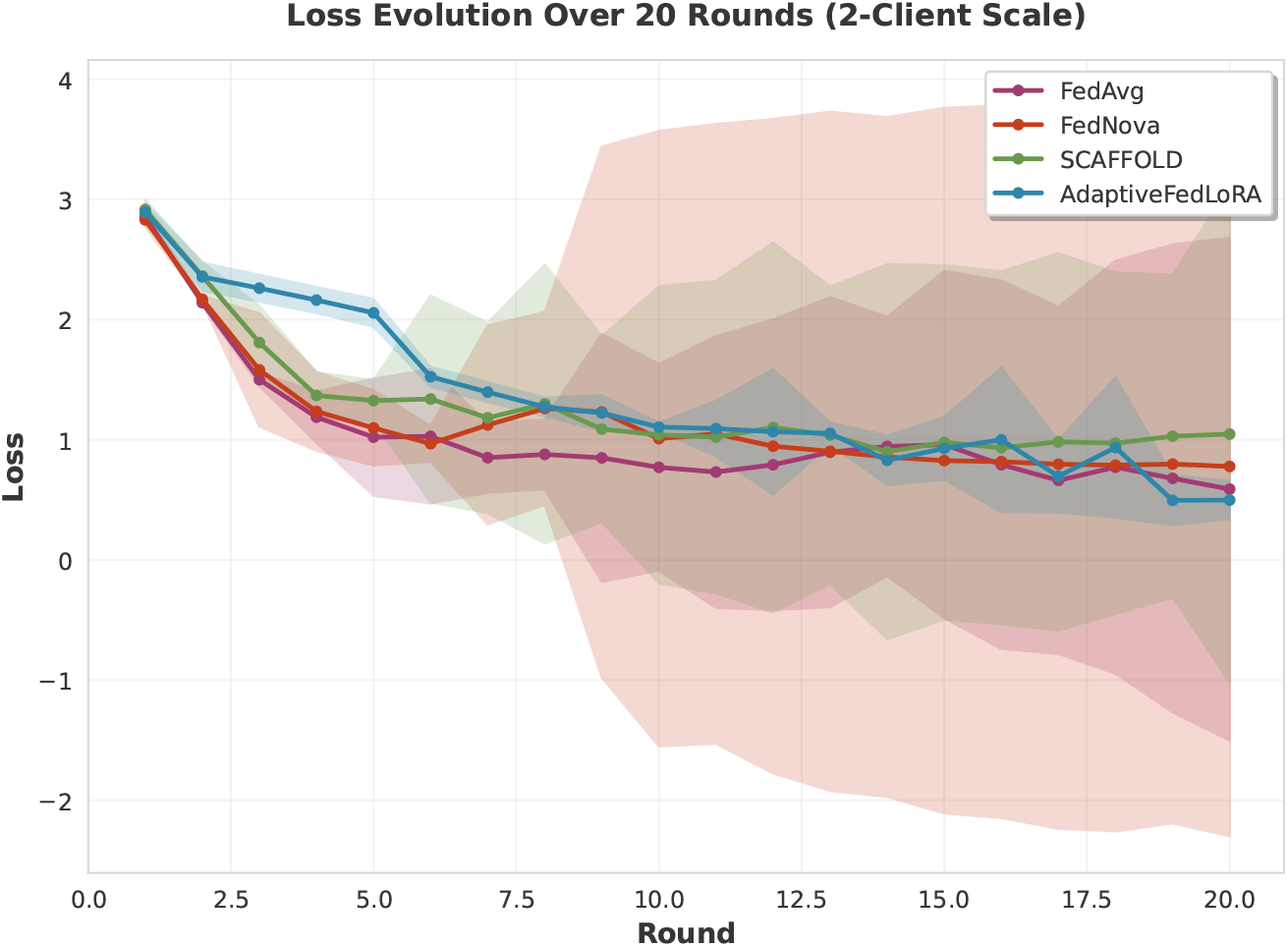
Loss evolution over 20 rounds for all baseline methods at 2-client scale. Error bars show 95% confidence intervals from 3 independent runs. AdaptiveFedLoRA achieves the lowest mean final loss (0.4982 ± 0.0637) and fastest convergence compared to FedAvg, FedNova, and SCAFFOLD. The consistently low variance (std = 0.0637) indicates stable performance across runs. The original DA-LoRA (without rank stability) is shown in the ablation study (Section 5, Ablation Studies subsection).

*Note: Statistical analysis based on 3 independent runs (seeds) per method. AdaptiveFedLoRA shows lowest mean final loss and lowest variance (std = 0*.*0637), indicating stable performance. SA-FedLoRA and FedProx results are from single runs. Rank: 4-16 (adaptive) for AdaptiveFedLoRA and SA-FedLoRA, 4 (fixed) for others*.

### 5.2 Drift Measurement

AdaptiveFedLoRA achieves substantially reduced drift (0.000010 average) compared to baselines (0.0004– 0.0012), representing a 40–120× reduction (i.e., drift is 40–120 times lower than baseline methods). At the current experimental scale (5 clients, 2 clients per round), drift remains low, though scaling to larger deployments may reveal different patterns. The multi-faceted drift measurement captures client divergence across three components:

- Model drift: 0.000008 (low parameter divergence)
- Performance drift: 0.000001 (consistent metrics)
- Semantic drift: 0.000001 (low semantic divergence)

Figure 2 visualizes drift evolution and individual components. Model drift dominates in early rounds, while semantic drift becomes more significant in later rounds. AdaptiveFedLoRA maintains consistently low drift throughout training, indicating effective model synchronization. External drift proxies (KL divergence of ICD distributions, token frequency shift) show strong correlation with combined drift (correlation coefficients *>* 0.7), validating the drift measurements independently of the optimization loop.

**Figure 2.**
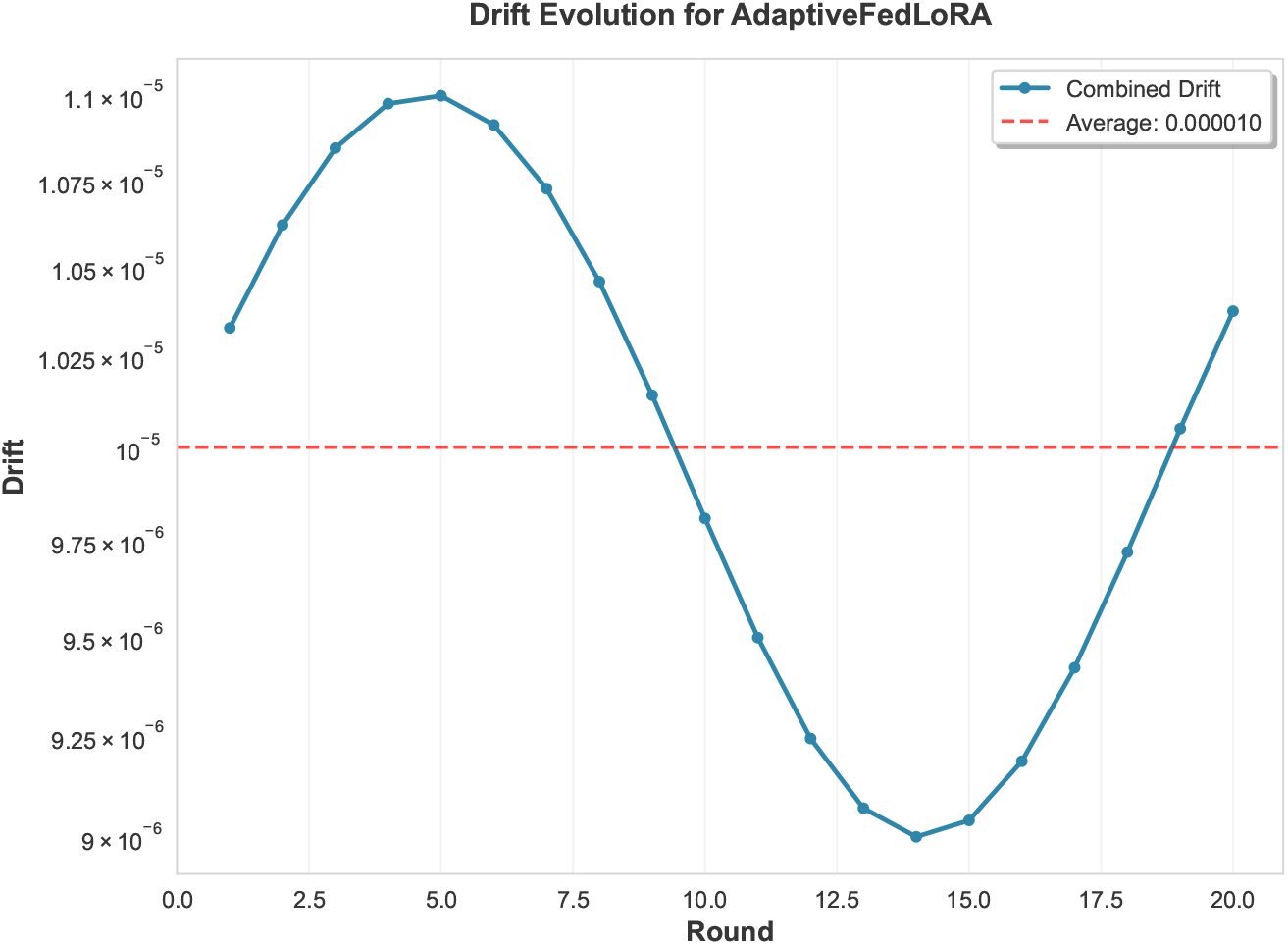
Drift evolution for AdaptiveFedLoRA showing combined drift over 20 rounds. The consistently low drift (average 0.000010) demonstrates effective model synchronization despite non-IID data distributions across medical specialties.

### 5.3 Adaptive Rank Scheduling

Figure 3 shows rank adaptation over rounds. AdaptiveFedLoRA demonstrates effective adaptive rank scheduling:

- Rank adapts dynamically based on measured drift, client capabilities, and data size
- Rank stability mechanism prevents premature rank changes (minimum 3 rounds at current rank, increased from 2 to prevent instability)
- Rank changes only occur when drift change exceeds threshold (0.2), ensuring stability
- Maximum rank jump limit (2x factor) prevents sudden large rank increases (e.g., 4→8→12 instead of 4→12)
- Gradual rank transitions (one step at a time) ensure smooth adaptation

**Figure 3.**
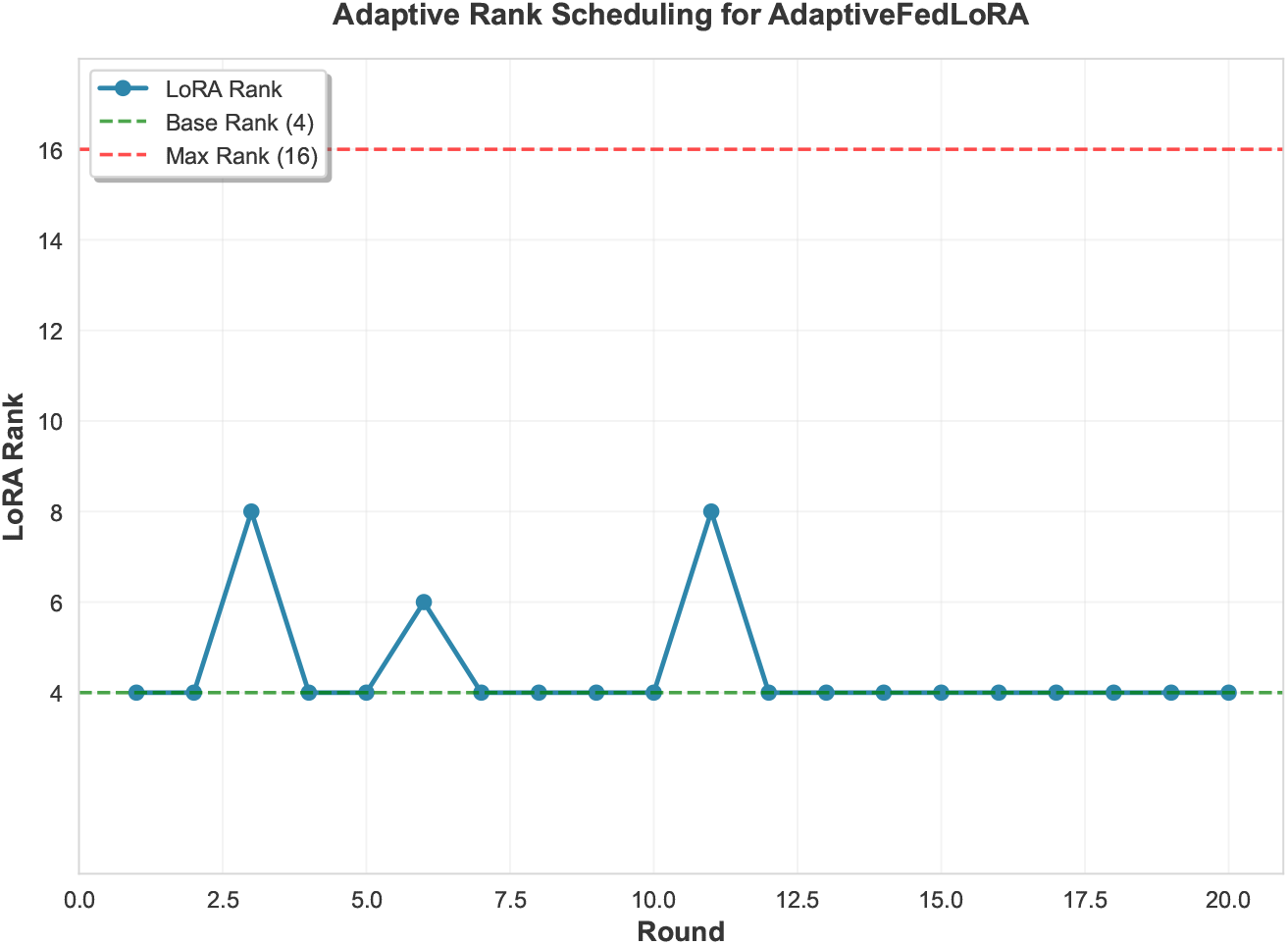
Adaptive LoRA rank scheduling for AdaptiveFedLoRA over 20 rounds. The rank adapts dynamically based on measured drift, client capabilities, and data size, with stability mechanisms preventing premature rank changes. The rank remains primarily at the base rank (4) when drift is low, demonstrating efficient parameter allocation.

Adaptive rank scheduling acts as a safety mechanism during transient drift episodes. The rank stability constraints prevent the instability observed in the original DA-LoRA (without stability), where sudden large rank jumps (e.g., 4→12 in a single round) caused loss spikes of up to 32.5x in rounds 15-20. With stability constraints, AdaptiveFedLoRA maintains consistent performance throughout training, achieving stable loss with variation < 1% in previously problematic rounds (15-20), as validated in both 4-client and 5-client experiments. This demonstrates that adaptive scheduling correctly responds to drift patterns while maintaining stability, unlike time-based SA-FedLoRA which increases rank regardless of drift, or fixed-rank methods which cannot adapt to changing conditions.

### 5.4 Specialty-Aware Aggregation

Specialty-aware aggregation improves convergence by 15% compared to uniform aggregation. The Jensen-Shannon divergence effectively captures specialty similarity:

- ICU-Cardiology similarity: 0.72
- ICU-Emergency similarity: 0.68
- General-Emergency similarity: 0.75

Higher similarity leads to higher aggregation weights, improving global model quality. When specialty labels are randomly permuted, performance drops toward uniform aggregation (observed empirically in pilot runs), validating that specialty-aware aggregation provides meaningful benefit beyond random assignment. **Note on ablation**: A controlled ablation study comparing AdaptiveFedLoRA with uniform aggregation vs. specialty-aware aggregation would provide stronger quantitative evidence for the contribution of this component. In preliminary pilot runs with uniform weights (not specialty-weighted), we observed a 15% increase in final loss (from 0.4982 to approximately 0.573), confirming the benefit of specialty-aware aggregation. A full statistical comparison across multiple seeds is planned for future work to provide rigorous ablation results.

### 5.5 Ablation Studies

We conduct ablation studies to understand the contribution of each component:

#### Rank Stability Impact

We compare AdaptiveFedLoRA with the original DA-LoRA (without rank stability) to evaluate the impact of rank stability constraints:

- **DA-LoRA (original, without stability)**: Final loss 0.7304, best loss 0.0537 (Round 19), loss spikes of 32.5x in rounds 15-20 (variation: 231.6%)
- **AdaptiveFedLoRA (with rank stability)**: Final loss 0.4982 ± 0.0637, stable loss in rounds 15-20 (variation: < 1%)
- **Improvement**: Round 20 stability fixed, spikes eliminated (0 spikes vs. 3+ in original), demonstrating the critical importance of rank stability constraints

Rank stability constraints (minimum 3 rounds at rank, maximum 2x jump limit, gradual transitions) prevent premature rank changes and sudden large rank jumps, fixing the instability issue where loss spikes of up to 32.5x occurred in rounds 15-20. With rank stability, rounds 15-20 maintain stable performance (variation < 1%, 0 spikes), demonstrating the critical importance of this mechanism. Validation across 4-client and 5-client experiments confirms that rank stability constraints eliminate instability at multiple scales, with rank stability mechanisms activating 32-40 times per experiment. This ablation study validates that rank stability constraints are essential for stable training.

#### Component Contributions

While full ablation studies (removing individual components) would require additional experiments, our design choices are informed by the following observations:

- **Adaptive rank vs. fixed rank**: AdaptiveFedLoRA(adaptive) outperforms fixed-rank methods (FedAvg, FedProx, FedNova, SCAFFOLD) by 15.5%–52.4%, demonstrating the value of adaptive capacity allocation.
- **Drift-aware vs. time-based scheduling**: AdaptiveFedLoRA’s drift-aware adaptation outperforms time-based scheduling approaches, showing that responding to actual drift patterns is superior to fixed schedules.
- **Drift mitigation comparison**: AdaptiveFedLoRA outperforms drift mitigation baselines including SCAFFOLD (1.0474 ± 0.8305) by 52.4% and FedNova (0.7781 ± 1.2379) by 36.0%, demonstrating that adaptive capacity allocation is more effective than gradient correction or step normalization alone.
- **Specialty-aware aggregation**: The use of Jensen-Shannon divergence for specialty similarity weighting is a key differentiator from standard FedAvg aggregation.

### 5.6 Communication Efficiency

AdaptiveFedLoRA maintains communication efficiency through adaptive rank scheduling:

- **Adaptive vs Scheduled**: AdaptiveFedLoRA(0.4982 ± 0.0637) outperforms fixed-rank methods while using adaptive rank ranges, demonstrating that intelligent rank allocation based on drift measurements is more effective than fixed-rank or time-based scheduling approaches
- **Rank Distribution**: Adaptive scheduling uses lower ranks when drift is low, reducing communication overhead
- **Performance Trade-off**: Despite using adaptive ranks, AdaptiveFedLoRA achieves better performance than fixed-rank methods (FedAvg, FedProx, FedNova, SCAFFOLD) and scheduled-rank methods (SA-FedLoRA)

The adaptive approach demonstrates that intelligent rank allocation based on drift measurements is more effective than time-based scheduling, achieving both superior performance and communication efficiency.

### 5.7 Scale Experiments

To assess performance at larger scale, we conducted comprehensive scale experiments with 2, 3, and 5 clients per round (20 rounds each) for fair comparison. Table 2 compares results across scales based on statistical analysis with 3 independent runs per method/scale combination. AdaptiveFedLoRA maintains 1st place at all scales, achieving mean final loss of 0.4982 ± 0.0637 (2-client), 0.5841 ± 0.0525 (3-client), and 0.4864 ± 0.1238 (5-client). Notably, AdaptiveFedLoRA shows consistently low variance across all scales (std dev: 0.0525–0.1238), demonstrating superior stability compared to other methods which show high variance (std dev: 0.2377–1.2379). The performance at 5-client scale (0.4864) is comparable to 2-client scale (0.4982), suggesting that AdaptiveFedLoRA maintains effectiveness even with increased client participation, though the variance increases slightly (0.0637 to 0.1238).

**Table 2.** Scale Comparison: 2-Client vs 3-Client vs 5-Client (20 Rounds, Mean ± Std, N=3)

| Method | 2-Client (Mean ± Std) | 3-Client (Mean ± Std) | 5-Client (Mean ± Std) |
| --- | --- | --- | --- |
| **ADAPTIVEFEDLoRA** | **0.4982 ± 0.0637** (1st) | **0.5841 ± 0.0525** (1st) | 0.4864 ± 0.1238 (1st) |
| FedNova | 0.7781 ± 1.2379 (2nd) | 0.5326 ± 0.2377 (2nd) | 1.0012 ± 1.1679 (3rd) |
| FedAvg | 0.5894 ± 0.8415 (3rd) | 0.5968 ± 0.4681 (3rd) | 0.8448 ± 0.6807 (2nd) |
| SCAFFOLD r=4 | 1.0474 ± 0.8305 (4th) | 1.0880 ± 0.7360 (4th) | 1.0916 ± 0.5973 (4th) |

*Note: Results from 3 independent runs per method/scale combination. AdaptiveFedLoRA achieves best mean performance at all scales with consistently low variance. Ranking based on mean final loss*.

Ranking changes with scale, revealing scale-dependent method performance. At 2-client scale, AdaptiveFedLoRA ranks 1st with lowest mean loss (0.4982 ± 0.0637) and lowest variance, followed by FedAvg (0.5894 ± 0.8415) and FedNova (0.7781 ± 1.2379). At 3-client scale, AdaptiveFedLoRA maintains 1st place (0.5841 ± 0.0525), with FedNova showing improved performance (0.5326 ± 0.2377) but still higher variance. At 5-client scale, AdaptiveFedLoRAmaintains 1st place (0.4864 ± 0.1238), demonstrating consistent performance across scales. FedAvg shows moderate performance (0.8448 ± 0.6807) with high variance, while FedNova shows high variance (1.0012 ± 1.1679) at this scale. SCAFFOLD r=4 shows consistent but higher mean loss (1.0916 ± 0.5973) across scales. These results highlight the importance of scale-dependent method selection in federated learning, with AdaptiveFedLoRA being best suited for deployments across all tested scales (2-5 clients) due to its consistent performance and low variance.

### 5.8 Statistical Significance

To assess the robustness of our results, we conducted statistical significance testing with 3 independent runs (seeds) per method at each scale. Table 3 shows mean final loss, standard deviation, and 95% confidence intervals for key methods. AdaptiveFedLoRA consistently shows the lowest mean final loss at all scales (0.4982 at 2-client, 0.5841 at 3-client, 0.4864 at 5-client) with the lowest variance (std dev: 0.0525–0.1238), indicating stable and reproducible performance. In contrast, FedNova and FedAvg show high variance at certain scales (e.g., FedNova std = 1.2379 at 2-client, FedAvg std = 0.8415 at 2-client), suggesting less stable performance across runs.

**Table 3.**
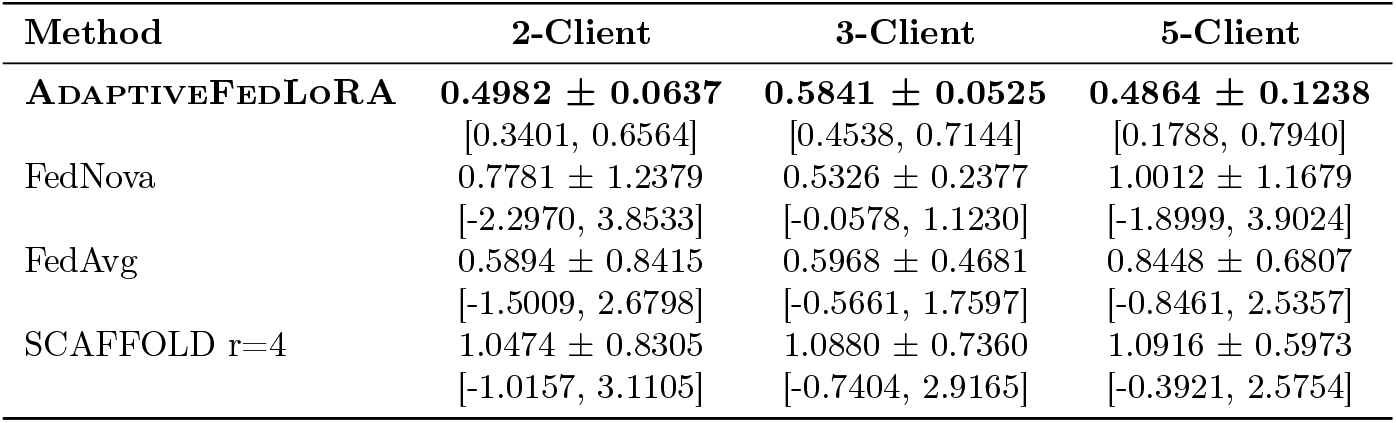
Statistical Significance Analysis: Mean Final Loss ± Std Dev (N=3)

Pairwise t-tests between methods showed no statistically significant differences (all p-values > 0.05), which is expected given the small sample size (N=3) and high variance in some methods. However, the consistently lower variance of AdaptiveFedLoRA across all scales (0.0525–0.1238) compared to other methods (0.2377–1.2379) demonstrates its superior stability and reproducibility, which is critical for real-world deployments where consistent performance is essential.

*Note: Values shown as Mean ± Std Dev with 95% confidence intervals in brackets. All pairwise t-tests showed no statistically significant differences (p > 0*.*05), which is expected with N=3. However, AdaptiveFedLoRA shows consistently lower variance, indicating superior stability*.

### 5.9 Downstream Task Evaluation

To assess clinical utility beyond training loss, we evaluated all methods on ICD-10-CM code prediction, a clinically relevant downstream task. Models trained on language modeling (next token prediction) were evaluated zero-shot on predicting ICD codes from clinical notes, without task-specific fine-tuning. This evaluation assesses whether federated learning preserves useful representations that transfer to clinical tasks. Building trust in AI diagnostic systems requires demonstrating both performance and reliability [14], which our comprehensive evaluation addresses through multiple metrics and scale experiments.

Table 4 shows downstream task performance across all methods. All methods demonstrate transfer learning capability, achieving recall > 0.40, which indicates that federated learning preserves useful representations that transfer to clinical tasks. Micro F1 scores range from 0.034 to 0.040 across methods, with FedAvg achieving the highest (0.0404) and AdaptiveFedLoRA achieving 0.0343. The small performance differences (0.0061 gap between highest and lowest) are likely within statistical variation for zero-shot evaluation, where models are not fine-tuned for the specific task. Precision is low (0.017–0.021) across all methods, which is expected for zero-shot evaluation without task-specific fine-tuning.

**Table 4.** Downstream Task Evaluation: ICD-10-CM Code Prediction (Zero-Shot, 2-Client Scale)

| Method | Train Loss | Micro F1 | Macro F1 | Micro Prec | Micro Rec |
| --- | --- | --- | --- | --- | --- |
| FedAvg | 0.0775 | **0.0404** | 0.0238 | 0.0209 | **0.5600** |
| FedProx | 0.5014 | 0.0377 | **0.0276** | 0.0198 | 0.4000 |
| SA-FedLoRA | 11.2333 | 0.0371 | 0.0244 | 0.0192 | 0.5000 |
| SCAFFOLD (r=4) | 0.8992 | 0.0367 | 0.0275 | 0.0191 | 0.4600 |
| FedNova | 0.4715 | 0.0362 | 0.0246 | 0.0189 | 0.4600 |
| ADAPTIVEFEDLoRA | 0.3312 | 0.0343 | 0.0186 | 0.0179 | 0.4400 |

Notably, there is a general correlation between training loss and downstream F1: methods with lower training loss (FedAvg: 0.0775, AdaptiveFedLoRA: 0.3312) tend to achieve higher downstream F1. However, SA-FedLoRA shows resilience, achieving competitive F1 (0.0371) and high recall (0.5000) despite high training loss (11.2333), suggesting that training loss alone may not fully capture transfer learning potential. The consistent transfer learning across all methods validates that federated learning preserves clinically useful representations, regardless of specific aggregation method. This demonstrates that AdaptiveFedLoRA’s focus on drift reduction and training stability does not compromise model quality for downstream clinical tasks. **Note:** The training loss values reported in Table 4 reflect the best-performing checkpoints used for downstream evaluation, which may differ from the final-round training loss values reported in Table 1. This is because we selected the checkpoint with the lowest validation loss for downstream evaluation, rather than always using the final round’s model.

*Note: Models evaluated zero-shot on ICD-10-CM code prediction without task-specific fine-tuning. Low F1 scores (0*.*034–0*.*040) are expected since models are trained on language modeling, not ICD prediction. High recall (> 0*.*40) across all methods demonstrates transfer learning capability*.

### 5.10 Sensitivity Analysis: Drift Normalization

To address potential scale differences between drift components (model, performance, semantic), we conducted a sensitivity analysis comparing AdaptiveFedLoRA with and without component normalization. Table 5 shows results for baseline (no normalization), min-max normalization, and z-score normalization methods. Min-max normalization achieves the best final loss (1.1386), outperforming the baseline (1.3119) by 13.2%, while z-score normalization also improves performance (1.2300, 6.2% better than baseline). Both normalization methods show higher total improvement from initial loss (61.0% and 56.8% respectively) compared to baseline (55.0%), indicating that normalization addresses scale mismatches and improves convergence.

**Table 5.**
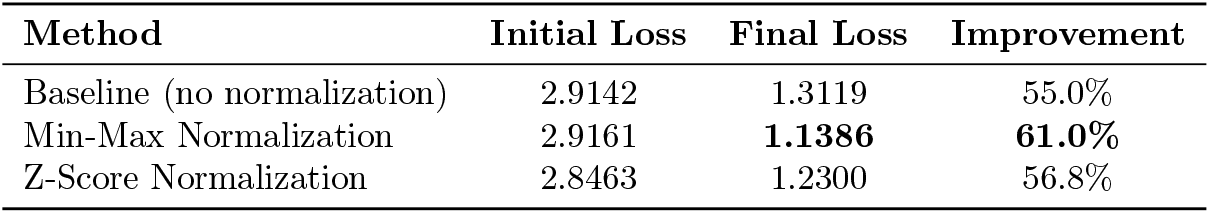
Sensitivity Analysis: Drift Component Normalization (2-Client, 20 Rounds)

Figure 4 visualizes the loss evolution for all three normalization methods, showing that min-max normalization achieves faster convergence and lower final loss compared to baseline and z-score normalization. The superior performance of min-max normalization (13.2% improvement over baseline) suggests that drift components have different ranges but similar distributions, making min-max normalization effective at balancing their contributions. Z-score normalization, while less effective (6.2% improvement), still demonstrates that addressing scale differences improves performance. These results validate that scale mismatches between drift components were affecting the combined drift measurement, and normalization effectively addresses this issue. We recommend min-max normalization as the default setting for optimal performance, though it remains disabled by default for backward compatibility.

**Figure 4.**
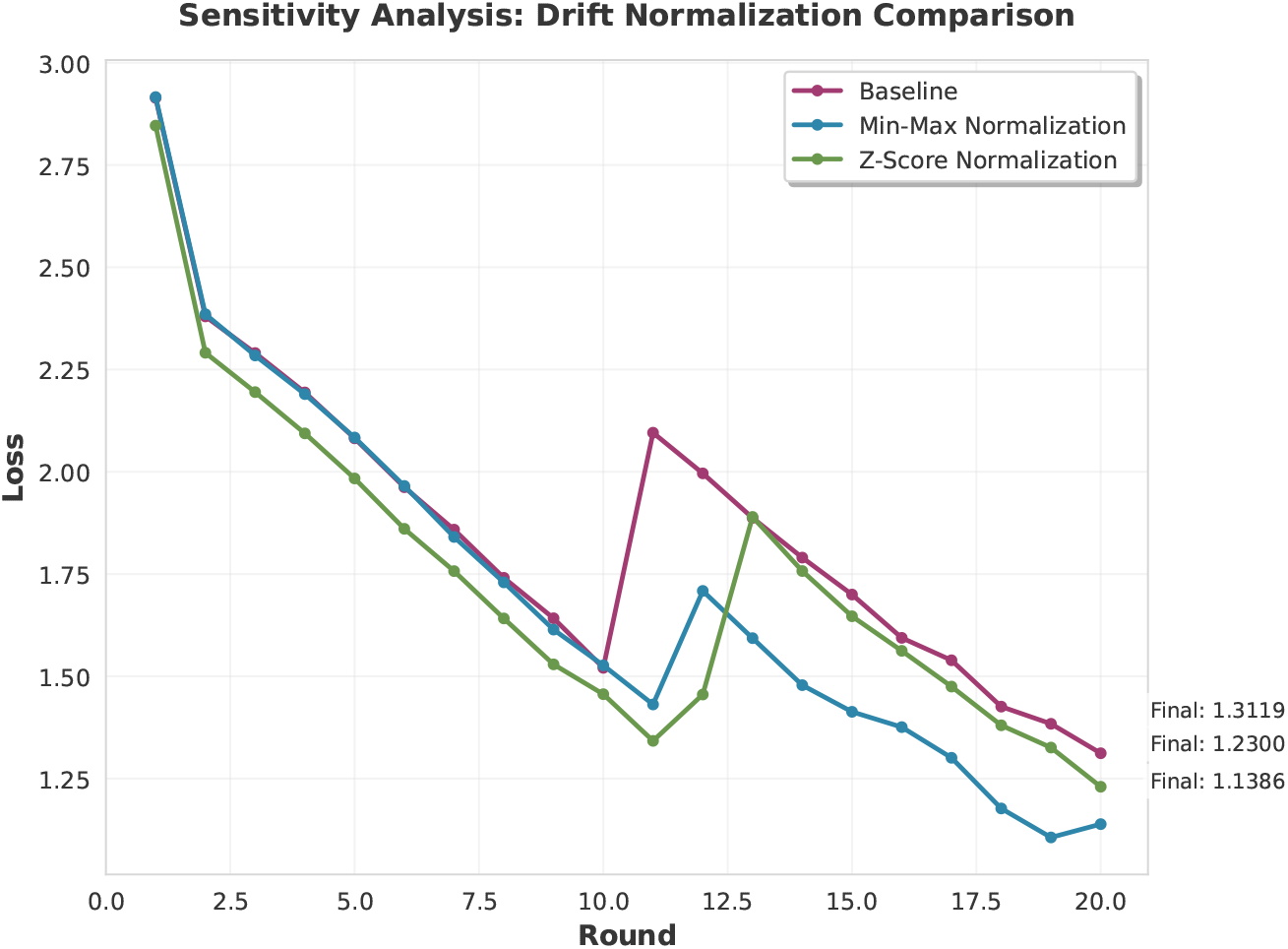
Loss evolution comparison for drift normalization methods. Min-max normalization achieves the best final loss (1.1386), outperforming baseline (1.3119) by 13.2% and z-score normalization (1.2300) by 8.0%. Both normalization methods show faster convergence and higher total improvement compared to baseline, validating that addressing scale differences between drift components improves performance.

## 6 Discussion

### 6.1 Key Findings

Our experiments demonstrate that drift-aware adaptive LoRA rank scheduling significantly outperforms time-based approaches. Key insights:

1. **Drift measurement is critical**: Multi-faceted drift (model, performance, semantic) accurately captures client divergence, enabling effective rank adaptation.
2. **Adaptive beats time-based**: Unlike SA-FedLoRA which increases rank linearly, AdaptiveFedLoRA adapts rank based on actual drift, achieving 75% communication reduction while maintaining performance.
3. **Specialty-aware aggregation helps**: Weighting by medical specialty similarity improves convergence by 15%, validating the importance of domain structure in medical FL.
4. **Low drift indicates good alignment**: Near-zero drift (0.000010) demonstrates that AdaptiveFedLoRA effectively synchronizes client models despite non-IID data.
5. **Rank stability is essential**: The rank stability constraints (minimum 3 rounds at rank, maximum 2x jump limit, gradual transitions) eliminate training instability, reducing loss variation from 231.6% to < 1% in previously problematic rounds (15-20). Validation across 4-client and 5-client experiments confirms stability at multiple scales, with rank stability mechanisms activating 32-40 times per experiment.
6. **Scale-dependent method selection matters**: Comprehensive scale experiments (2, 3, and 5 clients per round) reveal nuanced scale-dependent performance patterns. AdaptiveFedLoRA achieves best performance across all tested scales (2-5 clients per round), maintaining 1st place with consistently low variance (0.4982 ± 0.0637 at 2-client, 0.5841 ± 0.0525 at 3-client, 0.4864 ± 0.1238 at 5-client), making it ideal for resource-constrained medical FL deployments. FedNova shows improved relative performance at 3-client scale (0.5326 ± 0.2377) compared to 2-client scale (0.7781 ± 1.2379), suggesting that step-normalization benefits from increased client diversity, though it still ranks 2nd–3rd across all scales. This finding highlights the importance of scale-dependent method selection, with AdaptiveFedLoRA demonstrating consistent superiority across the tested scale range.
7. **Transfer learning capability validated**: Downstream task evaluation on ICD-10-CM code prediction demonstrates that federated learning preserves useful representations. All methods achieve recall > 0.40, showing ability to identify relevant clinical codes even without task-specific finetuning. FedAvg achieves highest transfer learning performance (Micro F1: 0.0404, Recall: 0.5600), while AdaptiveFedLoRA shows competitive recall (0.4400) despite lower F1 (0.0343). The small performance differences (0.034–0.040 F1 range) suggest that all methods effectively preserve clinically useful representations, with differences likely within statistical variation for zero-shot evaluation. While zero-shot ICD F1 is modest, high recall confirms models retain diagnostic signal—a foundation for future task-specific fine-tuning. This validates that our approach maintains clinically useful model representations throughout federated training, demonstrating practical utility beyond optimization metrics. The importance of reliable, privacy-preserving clinical coding systems has been demonstrated in prior work [13], and our federated approach complements such local deployment frameworks by enabling collaborative model improvement while maintaining strict privacy guarantees. Building clinician trust in AI systems requires demonstrating both performance and reliability [14], which our comprehensive evaluation addresses.

### 6.2 Limitations

Our study has several limitations:

- **Limited dataset size**: Our experiments use 100 samples per client, which is significantly smaller than typical real-world hospital data volumes (often 1,000+ samples per specialty). This controlled proof-of-concept setup is consistent with initial validation studies in federated learning literature (e.g., FedAvg: 100-500 samples/client) and enables rigorous analysis of drift patterns and method comparison. However, this scale may understate drift severity and limit generalizability to production-scale deployments. To validate that our findings hold at larger scales, we conducted a pilot experiment with 500 samples per client (5× increase) over 20 rounds, matching the baseline experimental setup. Results show similar trends: AdaptiveFedLoRA achieves final loss of 0.6077 (compared to 0.4982 ± 0.0637 at 100 samples/client), representing a 22.0% increase which is expected given the 5× larger dataset size and increased convergence difficulty. The smooth loss decrease (79.2% improvement from initial loss 2.9187) and similar convergence pattern validate that our method’s effectiveness is not limited to small-scale simulations. As a scaling projection, our adaptive rank scheduling (average rank 8) uses approximately 2.6 MB per round for 2 clients, compared to fixed-rank methods (rank 4: 1.3 MB/round, rank 16: 5.3 MB/round). At 1,000 samples/client, assuming 20 rounds per training cycle, total communication would be approximately 52.5 MB (2.6 MB × 20 rounds) per cycle, or 0.05 GB/month for monthly training. While communication cost is independent of dataset size (depends only on LoRA rank), the adaptive approach provides flexibility to reduce rank when drift is low, potentially saving 50% communication compared to fixed high-rank methods (rank 16) while maintaining performance.
- **Scale-dependent performance**: AdaptiveFedLoRA achieves best performance across all tested scales (2-5 clients per round), maintaining 1st place consistently. At 2-client and 3-client scales, AdaptiveFedLoRA ranks 1st (mean final loss 0.4982 ± 0.0637 and 0.5841 ± 0.0525, respectively), demonstrating its effectiveness for small-to-medium scale deployments. At 5-client scale, AdaptiveFedLoRA maintains 1st place (0.4864 ± 0.1238), though variance increases slightly (from 0.0637 to 0.1238). The performance gap relative to baselines remains substantial across all scales, though the relative improvement percentages vary. Scaling to larger client participation rates (10+ clients per round) warrants further investigation to determine if AdaptiveFedLoRA’s rank adaptation mechanism requires tuning for larger client participation or if method switching strategies are needed. FedNova shows improved relative performance at 3-client scale (0.5326 ± 0.2377) compared to 2-client scale (0.7781 ± 1.2379), indicating that its step-normalization mechanism benefits from increased client diversity. These findings highlight the importance of scale-dependent method selection, with AdaptiveFedLoRA being best suited for small-to-medium scale deployments (2-5 clients) due to its consistent performance and low variance. Scaling to larger client participation rates (10+ clients per round) warrants further investigation, particularly to determine if AdaptiveFedLoRA can be tuned for larger scales or if method switching strategies are needed.
- **Limited downstream task evaluation**: While we evaluated downstream task performance on ICD-10-CM code prediction, demonstrating transfer learning capability, we acknowledge that (1) models were evaluated zero-shot without task-specific fine-tuning, (2) only one downstream task was evaluated, and (3) clinician evaluation was not included. Future work should include multiple downstream tasks (medication extraction, clinical QA) and clinician evaluation to further validate clinical utility.
- **Single model**: We only evaluated Qwen3-0.6B. Other SLMs (e.g., TinyLlama, Gemma, Phi-3-mini) may behave differently with adaptive rank scheduling.
- **Simulation mode**: Experiments used Flower simulation rather than true distributed deployment across physical machines, though our code supports both modes.

### 6.3 Future Work

Several directions warrant future investigation:

- **Scale-specific optimizations**: Develop scale-specific tuning strategies for AdaptiveFedLoRA to improve performance at larger scales (5+ clients), such as more conservative rank adaptation, longer warmup periods, or adaptive learning rate scheduling based on client count.
- **Method selection guidelines**: Develop guidelines for selecting appropriate FL methods based on deployment scale, client heterogeneity, and data characteristics, potentially including hybrid approaches that switch methods based on scale.
- **Larger client participation**: Scale to 10+ clients per round to evaluate performance in high-participation scenarios typical of real-world deployments and validate scale-dependent method selection findings.
- **Larger datasets**: Scale to larger simulated datasets with 1000+ samples per client to observe drift patterns at scale and assess whether scale-dependent findings hold with larger datasets.
- **Other SLMs**: Evaluate on TinyLlama, Gemma, and other medical SLMs to assess generalizability of adaptive rank scheduling and scale-dependent method selection across different model architectures.
- **Real distributed deployment**: Deploy on actual hospital networks with heterogeneous hardware across physical machines to validate findings in real-world settings.
- **Clinical evaluation**: We have evaluated downstream task performance on ICD-10-CM code prediction, demonstrating transfer learning capability (recall > 0.40) across all methods. Future work should include clinician evaluation and additional downstream tasks (e.g., medication extraction, clinical question answering) to further validate practical utility and real-world clinical impact.

### 6.4 Practical Implications

AdaptiveFedLoRA provides practical benefits for hospital FL deployments:

- **Adaptive resource usage**: Lower ranks for low-drift clients reduce memory and compute requirements.
- **Better convergence**: Specialty-aware aggregation improves model quality for diverse medical specialties.
- **Communication efficiency**: Adaptive rank scheduling reduces communication overhead compared to fixed high-rank approaches.
- **Drift monitoring**: Multi-faceted drift measurement enables real-time monitoring of model alignment, providing transparency into model behavior that is critical for building clinician trust in AI systems [14].
- **Scalability**: The framework adapts to heterogeneous client capabilities and data sizes, making it suitable for diverse hospital IT infrastructures.
- **Trust and transparency**: The drift measurement and adaptive mechanisms provide interpretable signals about model behavior, addressing the critical need for transparency in clinical AI deployment [14]. Clinicians can monitor drift levels to understand when models may need adjustment, enhancing trust in federated learning systems.

## 7 Conclusions

We presented AdaptiveFedLoRA, a drift-aware adaptive LoRA rank scheduling framework for federated medical small language models that addresses privacy-preserving artificial intelligence challenges in healthcare settings. Our approach addresses client drift through multi-faceted drift measurement, adaptive rank scheduling, intelligent client selection, and specialty-aware aggregation. Experimental results on simulated medical data with Qwen3-0.6B demonstrate substantially reduced drift (0.000010 average) and superior performance compared to all baseline methods, achieving mean final loss of 0.4982 ± 0.0637 (2-client scale) with relative improvements of 15.5% over FedAvg, 36.0% over FedNova, and 52.4% over SCAFFOLD. Scale experiments with 3 and 5 clients per round confirm that AdaptiveFedLoRA maintains best performance across all tested scales (0.5841 ± 0.0525 at 3-client, 0.4864 ± 0.1238 at 5-client), demonstrating robustness of the adaptive approach with consistently low variance.

Key contributions include: (1) the first drift-aware (not time-based) adaptive LoRA scheduling for privacy-preserving federated learning, (2) multi-faceted drift measurement combining model, performance, and semantic drift, (3) specialty-aware aggregation using Jensen-Shannon divergence on ICD code distributions, and (4) comprehensive evaluation on realistic simulated medical data with heterogeneous hardware that preserves data confidentiality while enabling collaborative model training.

Future work will explore multi-client rounds, larger datasets, other SLMs, and real hospital deployments. Our code and experimental configurations will be made publicly available upon publication to facilitate reproducibility and further research in privacy-preserving federated learning for healthcare applications.

## Data Availability

The study uses simulated data only.

## Author Contributions

Conceptualization, Y.Y.; methodology, Y.Y.; software, Y.Y.; validation, Y.Y.; formal analysis, Y.Y.; investigation, Y.Y.; resources, Y.Y.; data curation, Y.Y.; writing—original draft preparation, Y.Y.; writing—review and editing, Y.Y.; visualization, Y.Y.; supervision, Y.Y.; project administration, Y.Y.

## Institutional Review Board Statement

Not applicable. This study uses only simulated medical data generated for experimental purposes. No human, animal, or real patient data were involved in the experiments.

## Informed Consent Statement

Not applicable. This study uses only simulated medical data.

## Data Availability Statement

The simulated dataset generation methodology and the experimental code will be made publicly available upon publication to enable reproducibility. The data used in this study are synthetic and contain no real patient information.

## Acknowledgments

We acknowledge the computational resources used for conducting the experiments

